# Nurse-physician Communication around Identifying Palliative Care Needs in Nursing Home Residents

**DOI:** 10.1101/2021.08.26.21262659

**Authors:** Jenny T. van der Steen, Esmée A. Jongen, Natashe Lemos Dekker, Lotje Bagchus, H. Roeline W. Pasman, Bregje D. Onwuteaka-Philipsen, Wilco P. Achterberg

**Author notes:** JTS and EAJ share equal first authorship. **Correspondence:** Jenny T. van der Steen, PhD, Leiden University Medical Center, Department of Public Health and Primary Care, Hippocratespad 21, Gebouw 3, P.O. Box 9600, 2300 RC Leiden, The Netherlands.

## Abstract

**Objective:** To assess experiences of medical practitioners who are on the staff of nursing homes with communication with nursing staff about identifying emerging and changing (palliative) care needs of residents of nursing homes in the Netherlands.

**Design:** Qualitative interview study.

**Setting and Participants:** Fifteen physicians and two nurse practitioners employed by eight care organizations in the western urbanized region of the Netherlands.

**Methods:** We conducted individual semi-structured interviews in 2018 informed by a topic list that was based on a qualitative dataset about facilitators to palliative care in dementia reported by elderly care physicians. The interviews were recorded, transcribed verbatim and analysed with Atlas.ti. We used both deductive and inductive coding adding refined codes related to communication.

**Results:** All interviewees expressed appreciation for nursing staff on whom they relied to communicate palliative care needs, yet they reported a variety of communication challenges around important changes in resident’s condition which were covered by two themes. (1) Teamwork was facilitated or impeded by team size and structure, quality of relationships and clarity in hierarchical relationships. (2) Continuity of information was affected by (in)effective routes of consultation and (lack of) detail in communicating observations.

**Conclusions and Implications:** Also in the case of on-staff physicians, functioning of the multidisciplinary team and accurate sharing of observed changes in nursing home residents’ condition are crucial for physicians to be able to address palliative care needs. The physicians’ expressions of how they would favor nursing staff to communicate can inspire interprofessional training, such as reporting objective observations and phrasing a clear request for help while avoiding overly demanding appeals.

## Introduction

Palliative care needs in nursing homes are projected to increase.^1,2^ Timely identifying changing resident’s care needs is crucial, whether physical, psychological, social or spiritual needs. Physicians are responsible for treatment decision-making, but nursing staff see residents more frequently and may be best positioned to identify changes.^3,4^ It is through conversation and consultation with nursing staff that physicians examine, present and interpret clinical data and decide on clinical actions, and nurses feel they realize best care.^5,6^

Ineffective interprofessional communication is a significant factor in medical error; nurse-physician communication is associated with patient safety and outcomes.^7,8^ Nurse-physician communication in end-of-life care has been reported as inadequate and impacting care.^6,9^ A classical ethnographic nursing home study indicated that it could cause unnecessary hospitalization.^10^ In Swedish nursing homes, the quality of nurse–physician communication related to the quality of pharmacological treatment.^11^

In the Netherlands, ‘elderly care’ physician is an officially recognized medical discipline with a 3-year training program.^12^ It includes training in social skills to communicate with all stakeholders and to participate in, and lead a multidisciplinary team. Further, these physicians, and increasingly also nurse practitioners (NPs) are employed by the nursing home and more present than physicians from community-based practices.^13^ We aimed to assess experiences of these on-staff medical practitioners regarding communication with nursing staff about identifying changing and emerging (palliative) care needs of residents.

## Methods

We conducted qualitative individual semi-structured interviews in the urbanized west of the Netherlands between October and December 2018. The Medical Ethical Committee of Leiden University Medical Center declared the study exempt from the Medical Research Involving Human Subjects Act (P17.256, 25 May 2018). The reporting adheres to the consolidated criteria for reporting qualitative studies (COREQ; available on request).

### Participants

Physicians and NPs practicing in nursing homes were invited to participate through nursing home administrations. With maximum variation sampling, we selected candidates based on diversity in gender, age, certification and the type of unit the participants practiced.

### Procedures and interview guide

After acquiring informed consent, individual interviews were conducted face-to-face in the nursing home by EAJ, female medical intern. A topic list (Appendix 1) addressed determinants of interprofessional communication reported in the literature.^7,9,14-18^ The list was also informed by studying a qualitative dataset with open-ended items from a survey among elderly care physicians on providing optimal palliative care with dementia.^19^ We started by asking about interdisciplinary communication in an open manner, continued with prompts from the topic list (e.g., mutual trust) and concluded asking what the physicians perceived as pleasant and unpleasant communication, and motivation to participate-to understand if this related to particular perceptions or experiences around nurse-physician communication. The interviewer made fieldnotes immediately following the interviews such as on perceived non-verbal hierarchical behavior.

### Analysis

Interview audio-recordings were transcribed verbatim (not returned) and anonymized. Due to recording failure mid-way, one-half interview was immediately described in detail from memory. The transcripts were coded based on recurring themes, using ATLAS.ti version 7.5.18 (2012). Coding was both deductive based on the literature and the qualitative dataset, and inductive, developing new codes that emerged from the interviews. Using the ‘framework method,’ a model for managing and mapping qualitative data systematically,^20^ we identified important themes regarding nurse-physician communication. A sample of two transcripts was analyzed independently from EAJ by NLD (anthropologist) and coding differences were discussed and resolved. During regular project team meetings with researchers with background in nursing, elderly care medicine, epidemiology, sociology and anthropology, intermediate findings were discussed. After 14 interviews, no new themes emerged. To increase confidence in the diversity of the sample related to the findings, we conducted three more interviews, with a very experienced elderly care physician and two non-specialized physicians.

## Results

Of 17 participants employed by 8 healthcare organizations, 14 (also) practiced on a psychogeriatric unit and most practiced on multiple types of units, totaling 29 (Table 1). The interviews lasted 36 to 62 (median 43) minutes. Initially asked in an open manner, the participants were quite appreciative about the communication with nursing staff concerning identifying residents’ (palliative) care needs. However, with specific probes, they raised a number of communication difficulties. Two main themes covering positive and negative experiences emerged from the data: teamwork, and continuity of information (Table 2).

**Table 1.** Characteristics of the interviewees (n=17); all employed by a nursing home organization to provide medical care

|  |  |
| --- | --- |
| **Gender** |  |
| men | 6 |
| women | 11 |
| **Profession** |  |
| elderly care physician | 12 |
| elderly care physician in training | 1 |
| medical doctor, no specialization | 2 |
| nurse practitioner | 2 |
| **Age, median (range)** | 49 (27-67) |
| **Years of experience in nursing home setting, median (range)** | 10 (0.6-30) |
| **Nursing home unit of practice (more possible)** |  |
| psychogeriatric unit | 14 |
| unit for chronic physical impairments | 7 |
| unit for rehabilitation | 4 |
| unit for palliative care | 1 |
| unit for intellectual disabilities | 2 |
| other: ambulatory care but reported on previous experience at psychogeriatric unit | 1 |

**Table 2.**
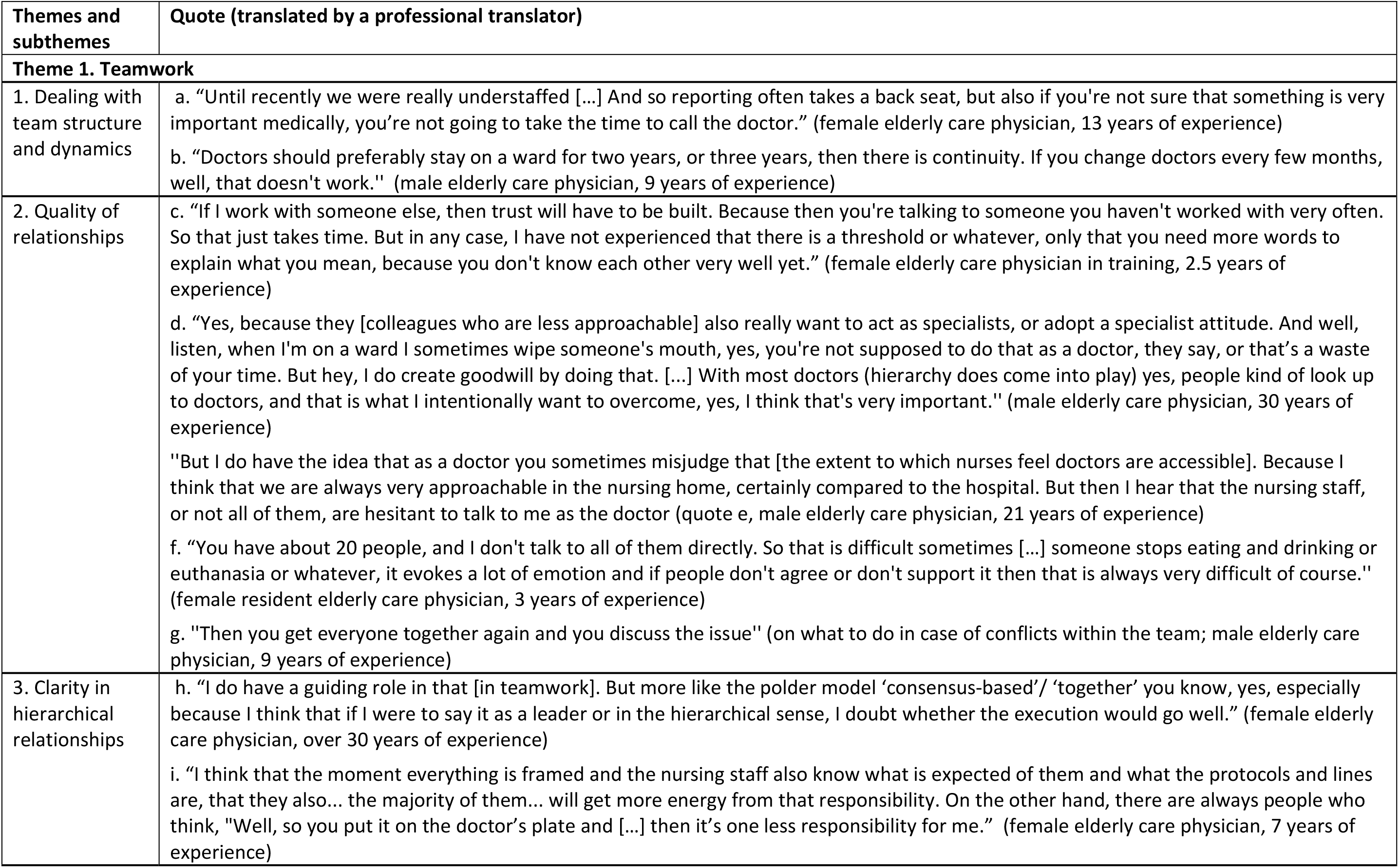
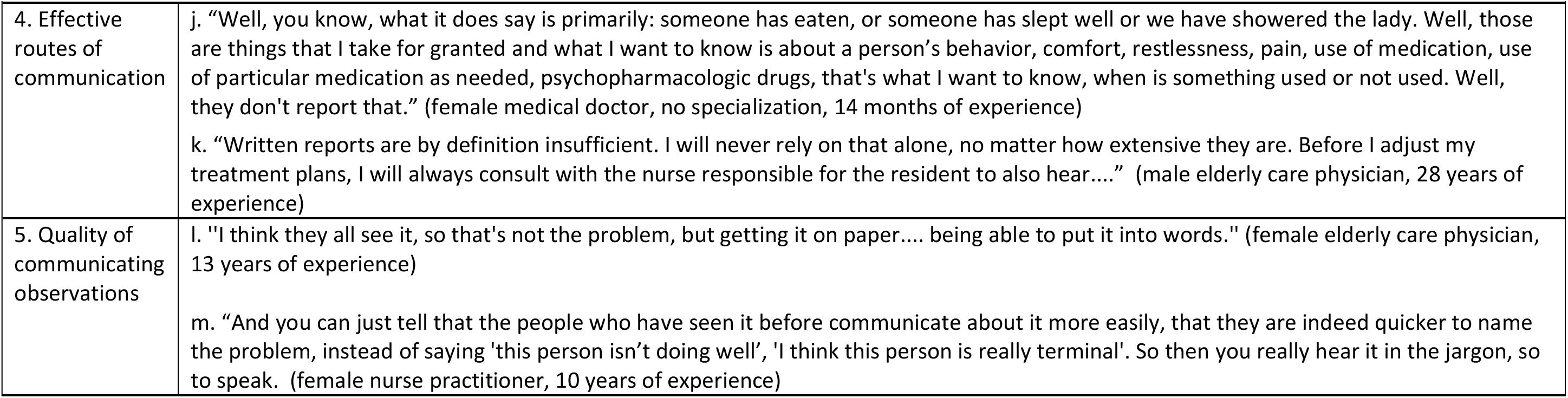
Themes, subthemes and exemplary quotes on communication with nursing staff

### Teamwork: Dealing with team structure and dynamics

Nursing staff mostly comprised nurse assistants and nurse aids. The physicians often did not know the exact size and structure of the care team they were working with. They regarded staff shortage, temporary staff, vacancies and high staff turnover as important barriers to communication on care needs. Staff shortage resulted in lack of time to communicate effectively including inadequate daily reporting (Table 2, quote a). Further, more acute matters would get priority and more gradual deterioration might go unnoticed or unreported. Lack of time was also believed to cause nursing staff to forget or stop following up physician’s orders, which caused frustration, less effective collaboration with physicians taking over to, for example, measure vital signs themselves.

Temporary workers to fill in vacancies would complicate communication because they would not know the residents, the care team and the physician. Temporary workers would not usually communicate with the physician. Also, they did not write reports when they were not familiar with the electronic record or were not authorized to access it. The physicians relied on team managers to assign experienced staff to work next to temporary workers. Some also noted the importance of continuity on part of the medical practitioners (quote b).

### Teamwork: Quality of relationships

Knowing each other both professionally and personally would lead to efficient communication through a good working relationship based on understanding and trust (quote c). This would be facilitated by continuity within the team and by regular contact with all staff including outside of scheduled meetings. Explaining reasons for decisions and involving nursing staff in the decision-making process enhanced trusting working relationships.

Interviewees mentioned a threshold for nursing staff to consult the physician due to perceived hierarchical difference (quotes d,e). Knowing each other may help, but, if the threshold was too low, nursing staff consulted the physician for minor issues.

Disagreements within teams or family complicated communication around identifying care needs as it posed emotional burden on nursing staff (quote f), especially when it concerned residents they had known for long or had grown particularly fond of. When residents deteriorated gradually, it would be important that nursing staff discussed changes with each other and report to the physician. However, not all teams had recurring meetings for such evaluations. Ideally, nursing staff was on the same page regarding residents’ condition prior to consulting the physician, but physicians felt they could also help settle disagreements (quote g).

### Teamwork: Clarity in hierarchical relationships

Clear leadership was seen as important by some physicians to make sure instructions such as performing regular measurements to detect gradual deterioration, are being followed up. Most physicians saw themselves as a supervisor rather than the leader (quote h), overseeing the multidisciplinary process and coaching the other disciplines if necessary. Some would see nursing staff to carry their own responsibilities, rather than being instructed by superiors, for them to perform their tasks with more dedication (with “more energy,” quote i), encouraging nursing staff in their role of identifying changes.

### Continuity of information: Effective routes of communication

The participants raised four routes of consultation to communicate identified changes: face-to-face, by phone, by electronic health record and by email. Face-to-face contact was seen as the most effective route and often sought in response to communication via other routes. It was the primary route to communicate gradual deterioration. Phoning was possible at any time and therefore suitable to communicate acute decline or when direct actions were needed, for example when residents were uncomfortable. Consultation via email was seen as less desirable considering privacy legislations.

Also, emails and written reports in the electronic record were likely to be missed by the physician. Some disclosed that they did not read the reports as incomplete or mostly for the nursing staff themselves (quotes a,j,k). Reporting on care goals was believed to be *un*helpful in this respect, because it would hinder reports on overall deterioration as this was not part of any care goal. To improve, the interviewees mentioned they explained the importance of objective and timely reporting on certain signals.

### Continuity of information: Quality of communicating observations

Communication on identifying (palliative) care needs was mentioned to be partially about translating a ‘gutfeeling’ into words (quote l), particularly concerning residents with dementia. The physicians felt that most nursing staff are aware of a resident’s deterioration, but have difficulty recognizing and explaining the exact problems, causing hesitancy to consult the physician. Usually the nurses with more education communicated with the physicians directly. However, they may have heard about changing resident needs second-hand.

The physicians saw it as their responsibility to clarify what staff says. Better clinical reasoning skills, understanding of medical jargon and language, staying calm and being clear in complex situations, having prepared through measurements, and a sense of longstanding involvement with the resident increased the quality of communicating observations. Experience was also helpful to interpret and translate what staff had seen before when residents were dying (quote m). Both education and experience promoted a pro-active attitude. Cultural or religious differences among staff in urban areas were seen as not much important. Rather, the overall team culture would be crucial in identifying and sharing concerns about resident’s changing care needs. Table 3 provides recommendations based on directly reported (un)pleasant communication or analyses of the interviews.

**Table 3.** Recommendations for Nurse-Physician Communication on Identifying (Palliative) Care Needs From the Perspective of Interviewed On-staff Physicians

| Physicians | Nursing staff | Both physicians and nursing staff |
| --- | --- | --- |
| Explain background when decisions need to be made and involve nursing staff<br>Encourage the nursing staff in their role<br>Explore (divergent) opinions and ensure you are accessible for communication on disagreement matters<br>Create a climate of mutual respect and trust<br>Create opportunities for regular and open contact with the members of the nursing staff<br>(also outside of scheduled meetings; *“just invest a lot when you are new somewhere, just drink many cups of coffee with staff when you are new and just show your face all the time. Even if you don't do anything, but just because of the fact, if they see your involvement, then communication is often OK as well”*; female elderly care physician in training, 2.5 years of experience). | Focus on clear communication clarifying the request about what help is needed while avoiding overly demanding appeals<br>Use objective arguments, substantiate with, for example recorded symptoms and measurements of vital functions, without your interpretations (e.g., as pain or anxiety)<br>Make sure you are well informed about a resident prior to consulting the physician<br>Use daily meetings within the nursing team to discuss a resident's condition and what should be discussed with the physician<br>Remain calm and confident when consulting the physician, also in more chaotic or acute situations. | Make sure all understand the background in case a decision should be made, and preferably decide in consensus<br>Discuss mutual expectations (telling the physician something *‘just so he/she knows’*, may be viewed as shifting responsibilities)<br>Plan regular multidisciplinary team meetings and audits<br>Get to know each other both professionally and personally. |

## Discussion

In this interview study, physicians on the staff of nursing homes appreciated nurses’ communication in their role as important team members yet reported a variety of communication problems around recognizing and communicating changes in residents’ condition and (palliative) care needs. Effective teamwork was facilitated or impeded by team structure and dynamics, quality of relationships and clarity in hierarchical relationships. Continuity of information was affected by routes of consultation and communicating observations. The physicians felt that nursing staff needs a pro-active attitude and good communication skills to make sense of, and report their observations-including if unrelated to a recorded care goal. Investing in relationships and educating nursing staff pay off when staff can be retained and can also help reduce staff turnover.

Physicians and nurses are trained differently which is highly relevant to identifying care needs. Nurses, but also elderly care physicians, are trained to view the patient from a holistic perspective. This is complex, systems-oriented and steeped in emotional intelligence. Physicians are generally trained to value an objective, more fact-based, structured and cognitive approach to patient care. They adopt succinct communication styles whereas nurses are trained to be highly descriptive.^21^ Nurses may start explaining patient’s background rather than with a clear question.^22^ Nurses often feel hurried by physicians,^7^ uninterested in what they say, feeling inferior.^6^ However, higher levels of information flow and self-organization are created by an open communication pattern between medical and nursing staff.^18^ Interprofessional education may improve physician-nurse communication,^21,23^ and tools structuring conversations.^22,24,25^

Face-to-face communication is an effective form of communication, yet this should be actively sought.^17^ Relational and informational continuity facilitate continuity of care.^26,27^ These were concerns even in the context of our study, with on-staff physicians supervising the team. The literature reports additional factors influencing nurse-physician communication in primary care and assisted living, from nurses’ or interprofessional perspectives, which we did not find, including territorialism, lack of time on part of the physician, difficulty reaching each other, not working with the same electronic records and unclear team goals.^7,9,14-18,28^

### Limitations and strengths

We examined views of physicians only. Physicians may perceive collaboration with nurses differently, or,^29^ more favorably than nurses do. Most interviewees were motivated by contributing to good palliative care rather than by particular experiences with physician-nurse communication. They might have had an interest in a positive account, aiming to recruit the interviewer to become an elderly care physician and fill vacancies. However, those aware of the medical student not yet enrolled in a specialization program conducting the interviews did not appear more positive.

## Conclusion and Implications

Despite physicians’ presence in nursing homes, there is room to improve communication with nursing staff on subtle changes in residents’ condition by systematic observation and communication along with creating opportunity for open communication. Future research may include ethnographic fieldwork to study nurse-physician encounters and trialing of tools to improve communication on palliative care needs and collaborative action.

## Data Availability

The qualitative interview data are not available as these cannot be anonymized adequately and we therefore did not ask permission from the interviewees to make the data FAIR.

## In Brief

Physicians who are on the staff of nursing homes felt that to identify residents’ palliative care needs, teamwork and effective consultation routes to communicate detailed observations are essential.

## Acknowledgements

We appreciate the willingness of the physicians to participate in the study and we thank them for their openness.

## Authorship

All authors were involved in the study’s concept and design. EAJ collected the data and EAJ and JTvdS drafted the article. EAJ and NLD conducted independent analyses while all authors were involved in synthesizing themes. JTvdS, NLD and WPA provided supervision. All authors were involved in interpretation of the data and all revised the article critically for important intellectual content and approved the final version to be published.

## Publication and presentation

This paper has been submitted for publication. An abstract of the work has been accepted for a presentation at the 17^th^ World Congress of the European Association for Palliative Care (EAPC), an online congress with live sessions 6 – 8 October 2021.

## Appendix 1 Topic List Interviews for Studying Physician-Nurse Communication around Identifying Emerging and Changing (Palliative) Care Needs in Nursing Home Residents in the Netherlands^7,9,14-19^

- Elderly care physician’s experiences with interdisciplinary teamwork in the nursing home setting in general.
- Elderly care physician’s experiences with interdisciplinary teamwork in palliative and terminal care in the nursing home setting.
- Accessibility
  - Contact moments
  - Routes of consultation
  - Availability of time
  - Physician on location
  - Points of improvement as regards to accessibility
- Transfer of information
  - Systems of transferring information
  - (Clarity of) written reports
  - Direct and indirect transfer of information
  - Performing tasks
  - Points of improvement as regards to accessibility
- Mutual trust
  - Familiarity
  - Working relationship
  - Conflicts
  - Points of improvement as regards to mutual trust
- Knowing each other’s role and responsibilities
  - Clarity of each other’s role and responsibilities
  - Value of role and responsibility.
  - Leadership
  - Autonomy and proactivity
  - Evaluation and giving feedback
  - Points of improvement as regards to knowing each other’s role and responsibilities
- Team
  - Structure and size of the team
  - Staff turn-over
  - Mutual goal
  - Points of improvement as regards to the team
- Personal factors
  - Emotional burden
  - Cultural factors
  - Language
  - Experience
  - Involvement
  - Knowledge and training
  - Competence
  - Points of improvement as regards to personal factors
- Experiences
  - Example of unpleasant communication and why this is experienced as unpleasant communication
  - Example of pleasant communication and why this is experienced as good communication
- Points that have not yet been discussed
- Motivation to participate

